# Construction of tongue image-based machine learning model for screening patients with gastric precancerous lesions

**DOI:** 10.1101/2023.01.10.23284379

**Authors:** Changzheng Ma, Peng Zhang, Shao Li

## Abstract

Screening patients with precancerous lesions of gastric cancer (PLGC) is important for gastric cancer prevention. It could improve the accuracy and convenience of PLGC screening to uncover and integrate valuable characteristics of noninvasive medical images involving in PLGC, by applying machine learning methodologies. In this study, based on unbiasedly uncovering potential associations between tongue image characteristics and PLGC and integrating gastric cancer-related canonical risk factors, including age, sex, Hp infection, we focused on tongue images and constructed a tongue image-based PLGC screening deep learning model (AITongue). Then, validation analysis on an independent cohort of 1,995 patients revealed the AITongue model could screen PLGC individuals with an AUC of 0.75, 10.3% higher than that of the model constructed with gastric cancer-related canonical risk factors. Of note, we investigated the value of the AITongue model in predicting PLGC risk by establishing a prospective PLGC follow-up cohort, reaching an AUC of 0.71. In addition, we have developed a smartphone-based App screening system to enhance the application convenience of the AITongue model in the natural population. Collectively, our study has demonstrated the value of tongue image characteristics in PLGC screening and risk prediction.

**Trial Registration:** ChiCTR2100044006

## Introduction

Gastric cancer is the second leading cause of cancer death in men and women in China, and more than 80% of patients are diagnosed at an advanced stage [1]. Patients with precancerous lesions of gastric cancer (PLGC), including intestinal metaplasia and dysplasia [2, 3], suffered a higher risk of gastric tumorigenesis, with an annual incidence of 0.25%-6% [4-6]. Screening and conducting reasonable health surveillance for patients with PLGC in the natural population would make a great contribution to facilitating the early prevention of gastric cancer.

Current screening methods suffered from some challenges, including invasive and relatively low accuracy, which limited their applications in population screening. On the one hand, although gastroscopy and biopsy are the gold standards for gastric disease diagnosis [7], it remains inefficient and unreasonable use with respect to gastric disease screening [8], resulting in approximately half of the patients screened with gastroscopy being non-atrophic gastritis and the early diagnosis rate of gastric cancer is less than 20%[1, 9]. On another hand, the application of serum markers that are commonly used as screening factors in various gastric cancer risk assessment methods, such as pepsinogen I/II and gastrin-17 [10-12], were limited by high sensitivity and specificity threshold for serological tests for risk screening in natural populations [13]. In addition, it remains cost-effective for both serum pepsinogen test screening and endoscopy, presenting difficulty for their practical application [14]. Thus, non-invasive and highly accurate screening models for PLGC patients are urgently needed.

As non-invasive indicators, tongue image characteristics were used for the surveillance of a broad spectrum of diseases, inspired by the diagnosis experience in traditional Chinese medicine [15-21]. Tongue image characteristics, including shape, color, and tongue coating, are believed to reflect the state of health and the progress and severity of disease, especially for digestive diseases as the tongue is physically connected to the digestive system. For example, massive studies indicated that tongue image characteristics have been shown to correlate with gastroscopy findings and can be used to predict gastric mucosal health [22, 23]. In addition, it was revealed that tongue surface and color characteristics as indicators can improve the accuracy of gastric cancer diagnosis [24]. From the pathologic perspective, the distribution of microorganisms on tongue coating has also been suggested to be related to the distribution in gastric tissue, which can be used as a marker for gastric disease risk screening [25-27]. Moreover, morphological markers based on tongue images are considered to be valuable for risk screening for other diseases, such as diabetes, fatty liver disease, and CoVID-19 [16, 28-31]. The above studies indicated that tongue image characteristics can assist in disease screening. Therefore, uncovering the risk characteristics of tongue images is potentially valuable for constructing PLGC screening models.

Recently, deep learning techniques are widely used in building biomedical image-based disease screening and prediction models [32-37]. For example, some studies have applied deep learning to predict cancer, such as prostate cancer and rectal cancer, based on medical images [34, 38]. For tongue images, some studies have used deep learning techniques to identify risk features in tongue images for the detection of diseases such as stomach cancer and diabetes [24, 39]. So deep learning techniques could be utilized to uncover tongue risk characteristics.

Therefore, to improve the efficiency of screening patients with PLGC, particularly in natural populations, in this study, we explored the tongue image characteristics of patients with PLGC and integrated them with traditional screening indicators to develop a PLGC screening model-AITongue. Then, we evaluated its screening effect by external validation in an independent dataset and explored its potential value as a risk predictor of PLGC in a follow-up dataset. We believed that our study would pave the way to address the urgent need for non-invasive PLGC screening in clinical practice.

## Methods

### Patient enrollment, and data collection

Gastritis patients were enrolled in this study at cChina-Japan Friendship Hospital and Yijishan Hospital of Wannan Medical College from 2015 to 2022. The experimental protocol was established according to the ethical guidelines of the “Declaration of Helsinki” and was approved by the Human Ethics Committee of Institution Review Board of Tsinghua University and the Chinese clinical trial registry. Inclusion criteria: Above 18 years of age with clear consciousness, clear language skills, and no barriers in communication and were willing to accept clinical investigation and sign informed consent. Exclusion criteria: Combined with heart, cerebrovascular, liver, kidney, hematopoietic system, and other primary diseases.

### Gastroscopy and histological examination

Using video endoscopes (Olympus Corp), upper gastroscopic examinations were performed by 2 gastroenterologists. Tissue samples for biopsy were reviewed blindly by 2 pathologists according to the criteria proposed by the Updated Sydney System and the Chinese Association of Gastric Cancer [40, 41]. The results of each biopsy were reported as normal, superficial gastritis, chronic atrophic gastritis, intestinal metaplasia, intraepithelial neoplasia, or gastric cancer, and each participant was assigned a global diagnosis based on the most severe gastric histologic finding among any biopsy. Helicobacter pylori (Hp) infection status was determined by enzyme-linked immunosorbent assay for plasma IgG [42].

### Data pre-processing and structuring

Python (3.7.0) and PyTorch was used for tongue image preprocessing. We trained a tongue detection model using the YOLOv5 deep learning model [43]. With the model tongue images were detected and cut into tongue body images for subsequent analysis.

Basic information and clinical symptom characteristics were obtained from electronic medical records. The content included baseline information (gender, age) and symptom characteristics (xerostomia, bitter taste, gastric distention, stomach pain, etc.). All the above indicators were structured as two-category labeled data. Among them, age was divided into >50 and <=50 years based on the differentiation effect. Multiple interpolation methods were used to fill in the missing data. Tongue labels (fissure, etc.) were labeled by physicians.

### Screening model construction

The main body of the image classification part of AITongue model is the ResNet50 model [44]. ResNet50 is a commonly used model in deep learning and is heavily used in research on images. Since this model introduces the residual block, it has a wide range of applications and good performance in the field of image classification. The residual blocks in the ResNet50 model are structured as two bottlenecks (BTNK), which are BTNK 1, BTNK 2 and their structure diagram is in Appendix 1. CONV is the convolution block, BN is the batch normalization block, and Relu is the activation function in the bottleneck.

The tongue image was reshaped as 224*224*3 and input to the model. Then, after the ResNet50 module, an MLP and a sigmoid module were used to classify the tongue images into two categories: high-risk and low-risk. Next, the image category labels were fed into a logistic regression model along with labels such as basic information. Finally, the model output the predicted scores for PLGC.

### Statistical Analysis

All analysis procedures were performed using Python (3.7.0) and the sklearn package. Tongue diagnostic labels (TDL) and clinical symptoms with statistical significance (p<0.05) by univariate and multivariate analyses were included in the model. Binary logistic regression was used to construct screening models. Chi-square tests were applied to calculate the significance of independent variables for PLGC. Pearson’s correlation coefficient was applied to evaluate the correlation between independent variables. Accuracy, sensitivity, specificity, recall, precision, ROC curve, and AUC were used as evaluation metrics to evaluate model performance.

## Results

### The overall design of our study

All patients underwent gastroscopy and pathology and were divided into three cohorts: development cohort, validation cohort, and follow-up cohort for model development, external validation, and risk prediction, respectively, where two categories, including PLGC and non-PLGC, were classified for patients based on pathological diagnosis (Table 1). In detail, we developed AITongue model for PLGC screening and did internal validation on the development cohort, which had a total of 325 patients, including 55 PLGC patients and 270 non-PLGC ones. Then, we performed external validation on the validation cohort, which had a total of 1995 patients, including 171 PLGC patients and 1824 non-PLGC ones. Of note, we evaluated AITongue model for risk prediction of PLGC on the follow-up cohort, in which only non-PLGC patients were enrolled in the baseline, and patients with PLGC and non-PLGC in the endpoint were classified as Pro and non-Pro, respectively, after a mean follow-up time of 22 months (Figure 1).

**Table 1.**
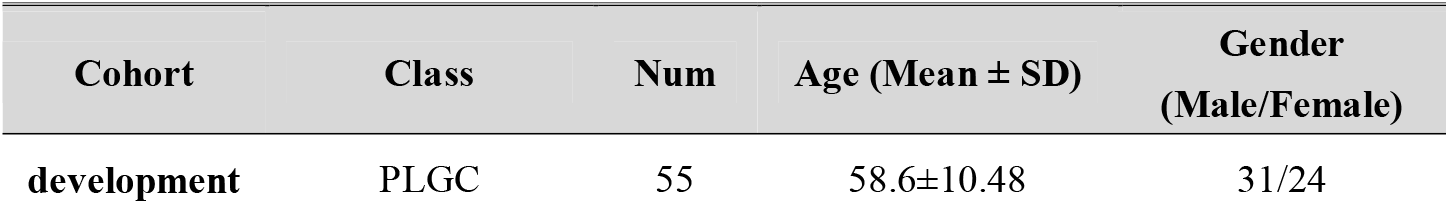
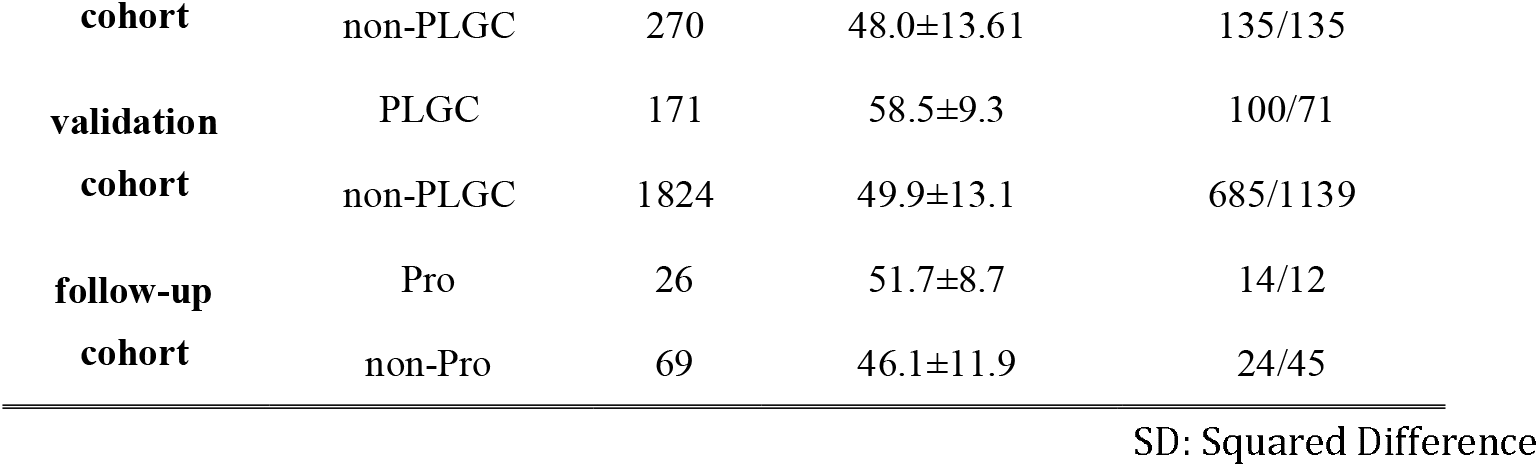
Basic information of 3 Cohorts.

**Figure 1.**
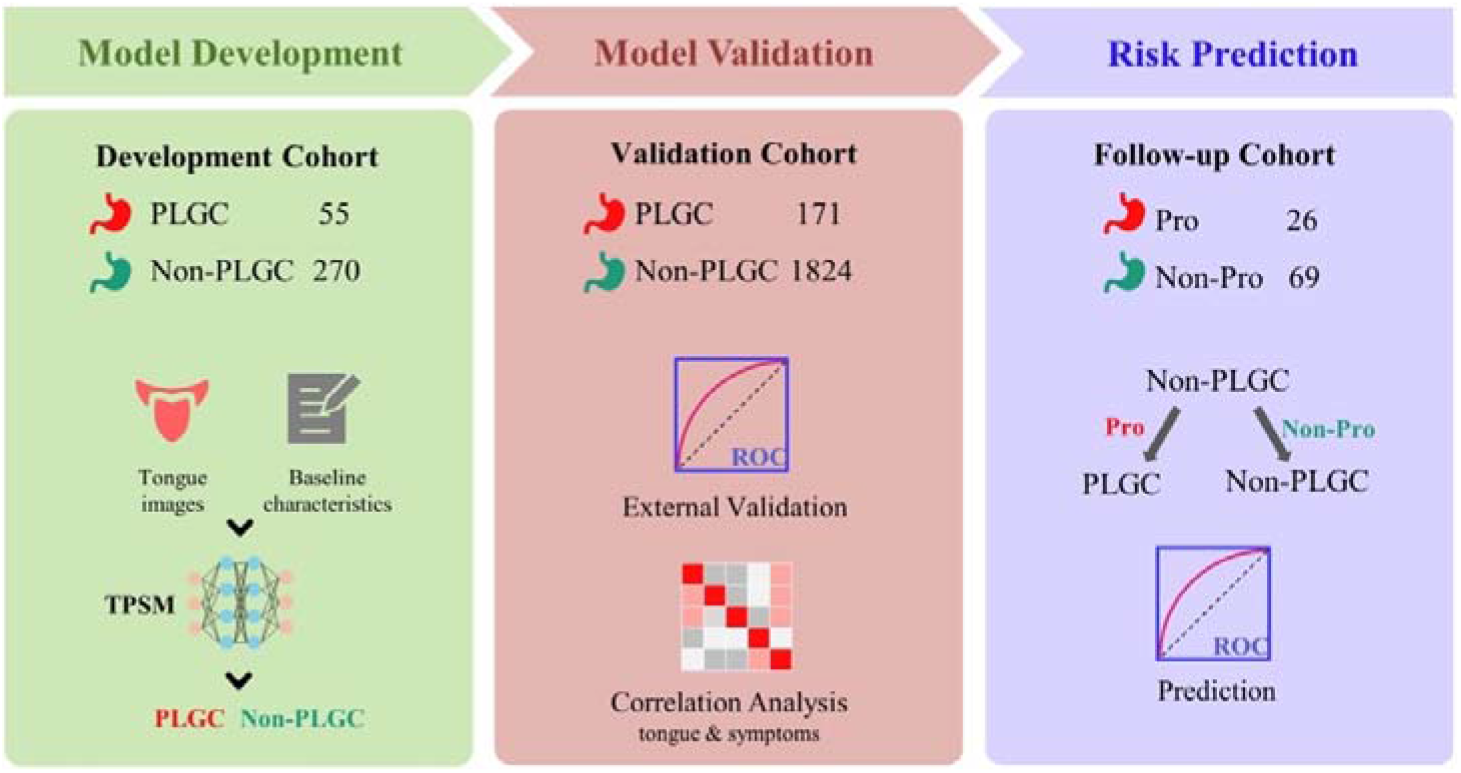
The study outline of work flow.

AITongue model was constructed based on deep learning and was shown in Figure 2b. The model took the preprocessed tongue images as input and output the risk score of the images for PLGC screening. The preprocessed tongue images were shown in Figure 2a. And then, the risk scores of the images were input to a logistic regression model together with baseline labels, and finally, the PLGC classification results were output.

**Figure 2.**
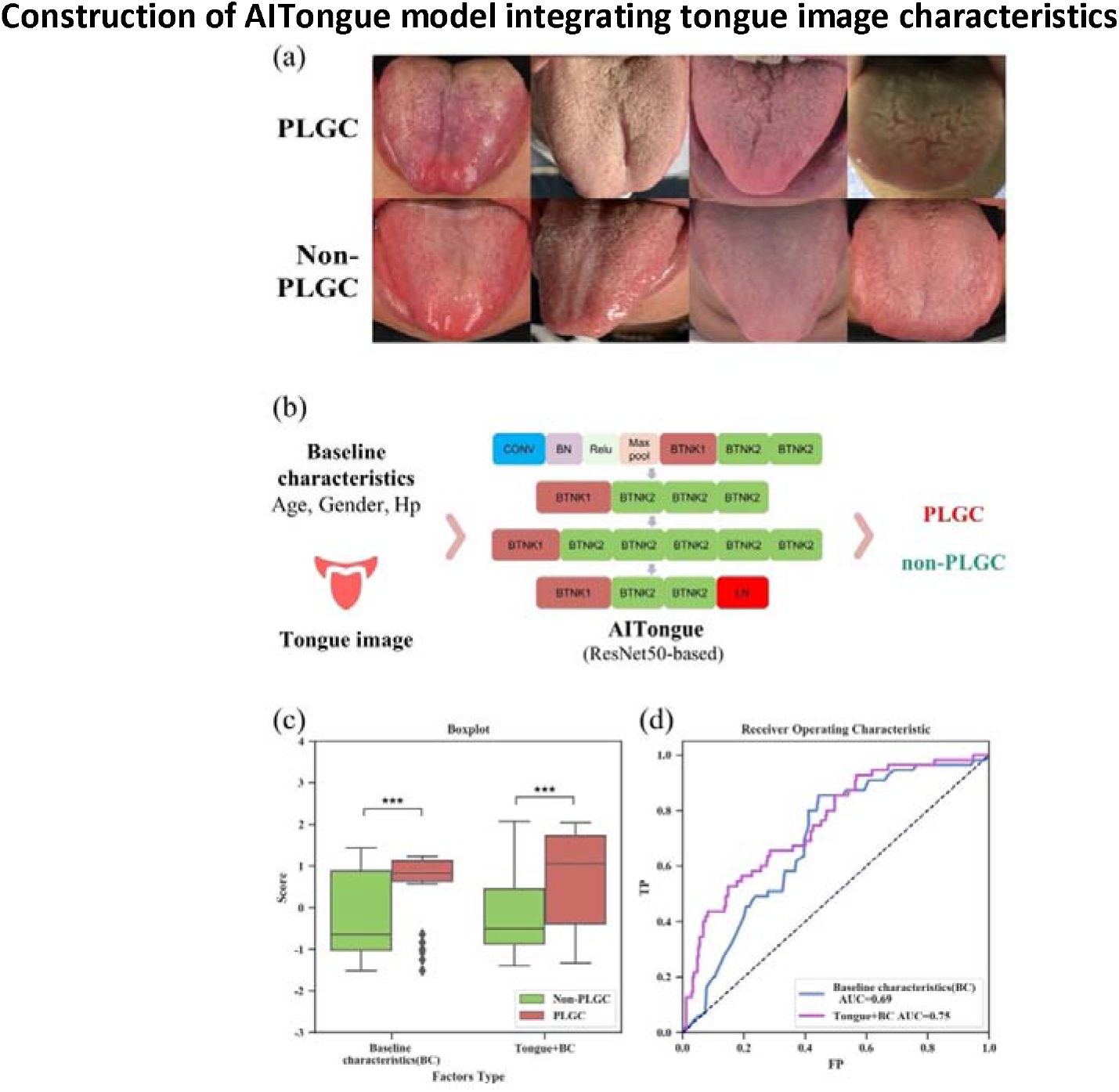
Construction of AITongue model and results for PLGC screening based on tongue images. A. Example of tongue images of PLGC and non-PLGC. B. ResNet50-based deep learning screening model-AITongue. C. Boxplot of classification score comparisons with and without the inclusion of tongue images for PLGC screening. D. ROC curves and AUC comparisons for PLGC screening. (***: p<0.001)

The inclusion of tongue image characteristics improved the PLGC screening effect by 8.7%. The development cohort was used as training data, and the screening model was validated internally by a five-fold cross-validation method. The boxplot showed that the classification scoring results with the introduction of tongue image were more discriminative for PLGC and non-PLGC (Figure 2c). The screening model with baseline (age, sex, Hp) as input had an accuracy of 0.60, a sensitivity of 0.80, a specificity of 0.56, and an AUC of 0.69. With the introduction of tongue images, the accuracy was 0.69, sensitivity was 0.71, specificity was 0.69, and AUC was 0.75. In terms of AUC, the screening effect was improved by 8.7% (Figure 2d). Therefore, it indicated that the introduction of tongue image characteristics contributed to the efficiency of PLGC screening.

The medical significance of risk features in the tongue images was interpreted and a risk typing analysis of the tongue images was performed. Traditional Chinese medicine (TCM) practitioners labeled 325 tongue images based on the tongue diagnosis method. The above screening model classified 94 and 233 tongue images as high-risk and low-risk, respectively. We found that 5 of the tongue diagnosis labels were statistically significant (p<0.05), namely greasy, fissure, dark, coating (yellow), and coating (thick), which explained, to some extent, the medical significance of the risk features in tongue images and suggested that there may be some value of TDL for PLGC screening as well (Table 2).

**Table 2.** TDL analysis of high and low-risk tongue images classified by AITongue model.

| Characteristics | High Risk (94) |  | Low Risk (233) |  | P value |
| --- | --- | --- | --- | --- | --- |
|  | Num | Rate | Num | Rate |  |
| Coating (Yellow) | 16 | 0.17 | 16 | 0.07 | 9.6e-3 |
| Greasy | 28 | 0.30 | 34 | 0.15 | 2.6e-3 |
| Fissure | 19 | 0.20 | 26 | 0.11 | 4.8e-2 |
| Coating (Thick) | 14 | 0.15 | 15 | 0.06 | 2.6e-2 |
| Dark | 12 | 0.13 | 7 | 0.03 | 1.6e-3 |

### External validation for PLGC screening

To validate the AITongue model and analyze the interpretability of tongue image characteristics in the AITongue model, the tongue images in the validation cohort were transformed into interpretable TDL by TCM tongue diagnosis, to directly quantify the role of different features of tongue images in PLGC screening [45, 46].

TDL that were statistically significant for PLGC were screened as indicators before validation. Gender, age, and 5 TDL showed significance in univariate and multivariate analyses of the validation cohort (Table 3). TDL such as teeth marks that did not show significance were in Appendix 2. The 5 significant characteristics were greasy, fissure, dark, coating (yellow), and coating (thick), which were included in the screening model to validate the effect of introducing tongue image characteristics to enhance PLGC screening.

**Table 3.**
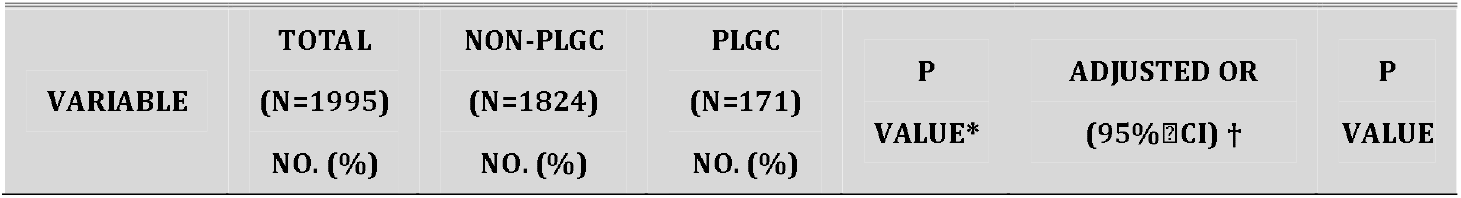
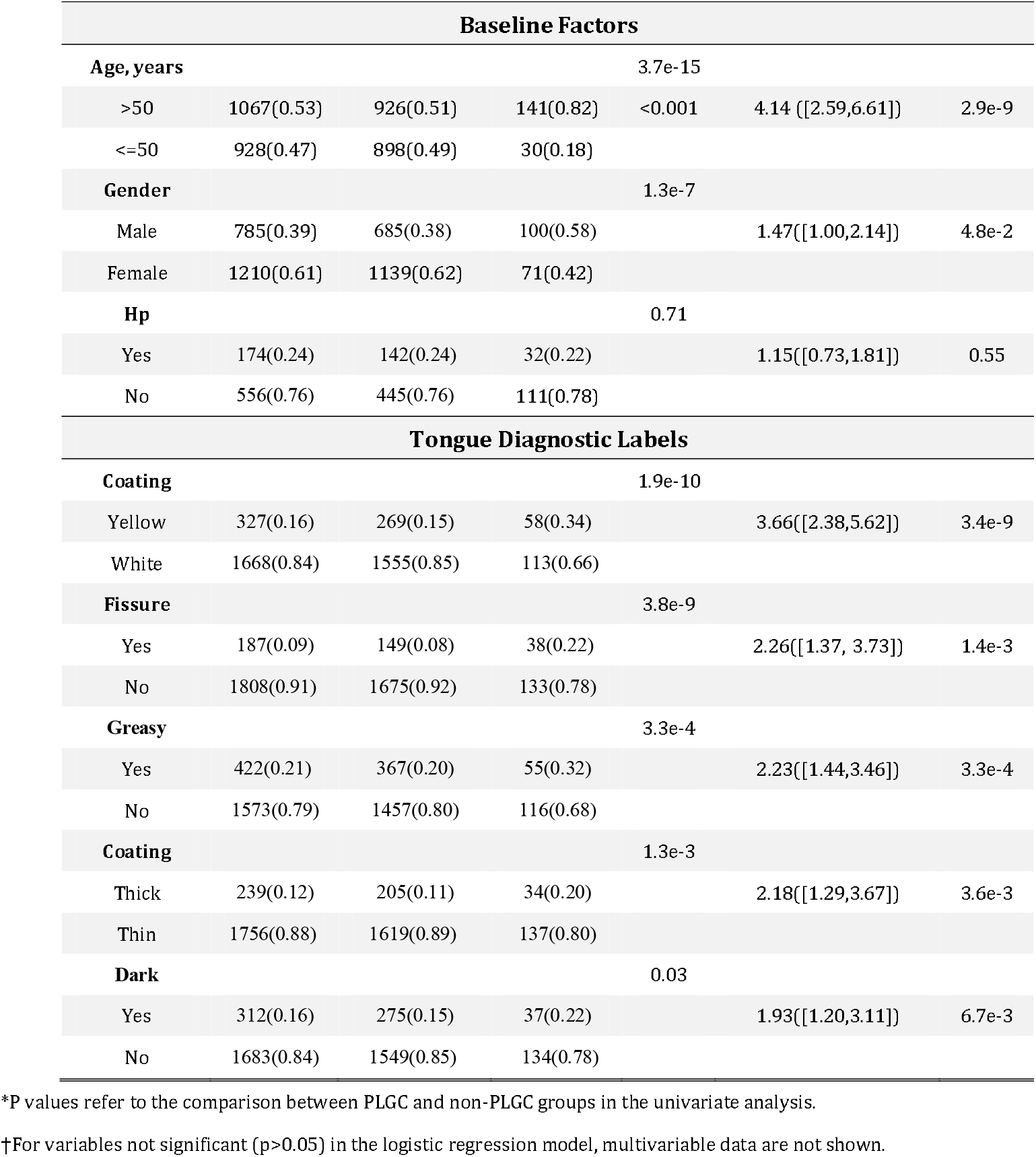
Univariate and multivariate analysis of baseline factors and TDL in PLGC screening.

The effectiveness of PLGC screening was improved by 10.3% in the validation cohort. We used a validation cohort to validate the effectiveness of tongue image characteristics for PLGC screening by logistic regression. The boxplot showed that the classification scoring of introducing tongue image characteristics was more discriminative for PLGC and non-PLGC (Figure 4). The accuracy of the baseline screening model with baseline (age, sex, Hp) as input was 0.53, sensitivity was 0.76, specificity was 0.51, and AUC was 0.68. With the introduction of TDL, the accuracy was 0.64, sensitivity was 0.72, specificity was 0.63, and AUC was 0.75. In terms of AUC, the screening effect was significantly improved by 10.3% (p<0.01) (Figure 3). The results illustrated that the introduction of TDL helped to improve the screening efficiency of PLGC and validated the effectiveness of tongue image characteristics for PLGC screening.

**Figure 3.**
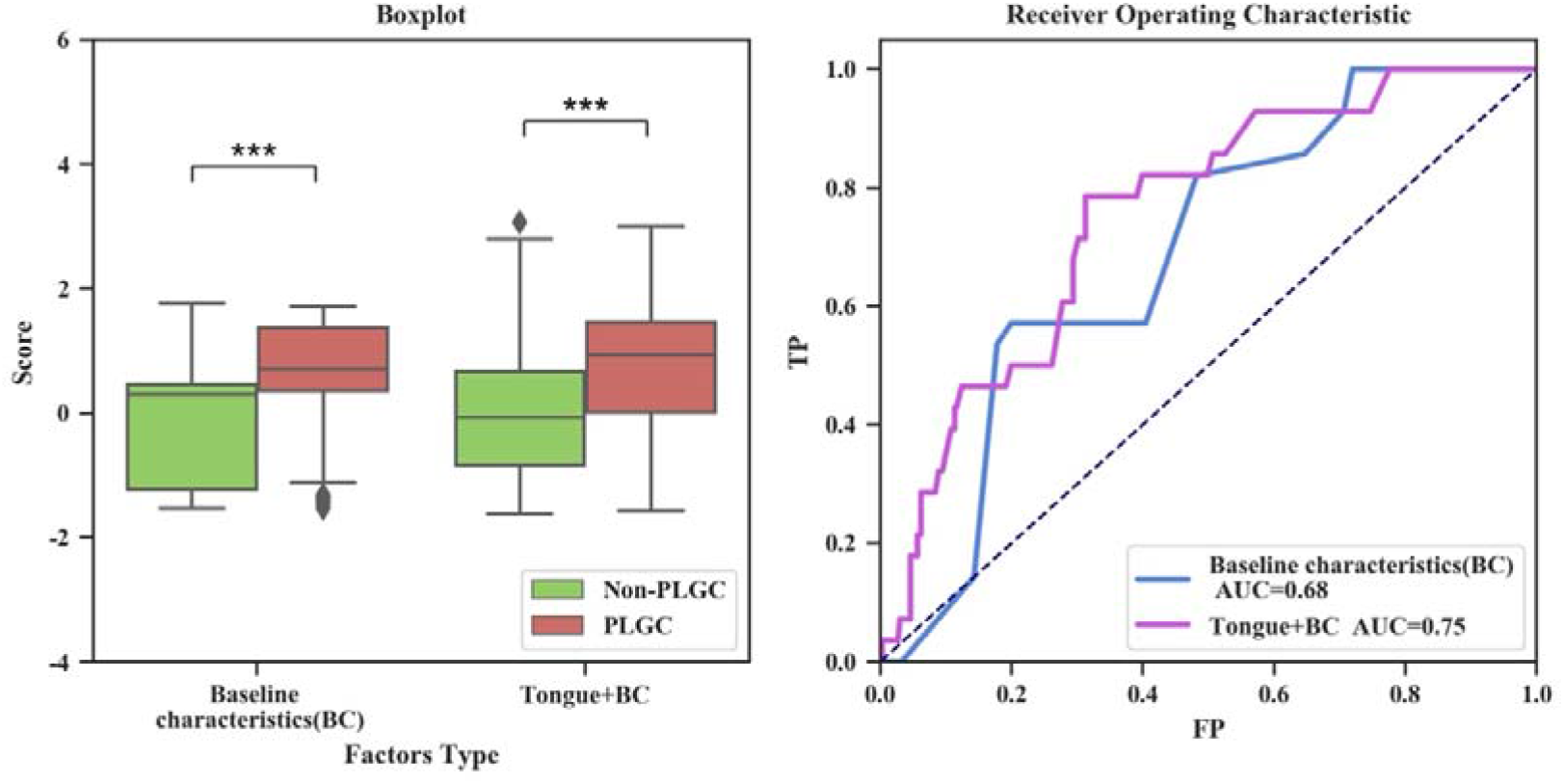
Boxplot of screening score and ROC curves comparisons with and without the inclusion of tongue image characteristics for PLGC screening. (***: p<0.001)

**Figure 4.**
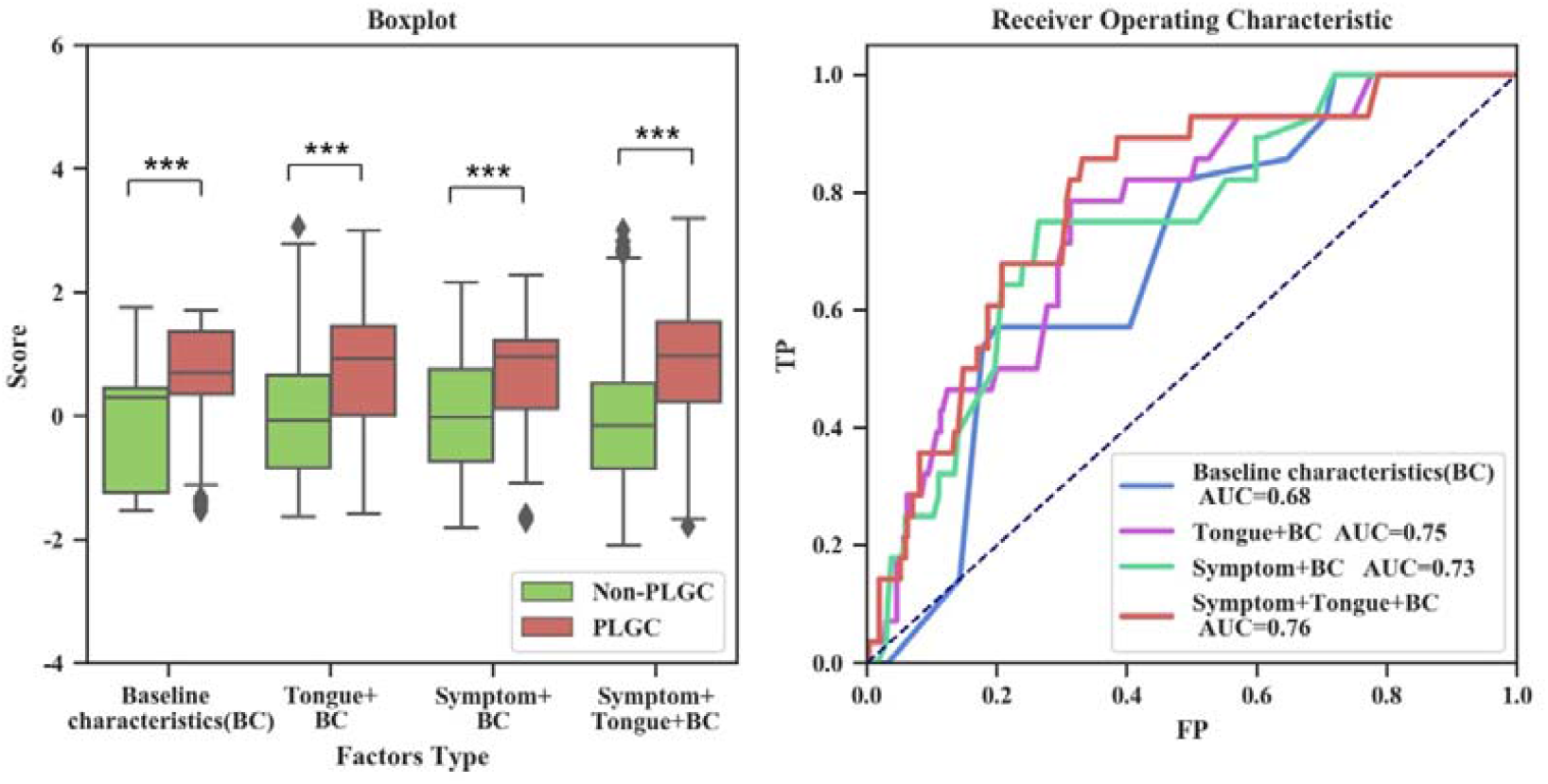
Boxplot of screening score and ROC curves comparisons with and without the inclusion of symptoms factors for PLGC screening. (***: p<0.001)

In addition, the clinical symptom characteristics of patients were counted in the validation cohort to assess their correlation with tongue image characteristics and their effect on screening.

First, we investigated the correlation between clinical symptoms and PLGC. The 3 symptom factors (xerostomia, bitter taste, belching) showed significance in univariate and multivariate analysis and they were included in the screening model to validate the effect of introducing clinical symptoms characteristics to enhance PLGC screening (Appendix 2). Factors that did not show significance such as stomach pain, bloating, chilliness, and loose stools were in Appendix 3. Further, we incorporated symptom characteristics into the screening model to evaluate the enhancement effect of introducing symptom characteristics for PLGC screening. The validation cohort was used as training data to construct a logistic regression model and performed five-fold cross-validation. The boxplot showed that the classification scoring result after introducing symptom characteristics has a small improvement in the discrimination between PLGC and non-PLGC (Figure 4). The AUC of the baseline screening model with baseline (age, sex, Hp) as input was 0.68. The AUC with tongue label and baseline as input was 0.75. The AUC with clinical symptoms and baseline as input was 0.73. The AUC with TDL, clinical symptoms, and baseline as input was 0.76 (Figure 5). The results indicated that the introduction of clinical symptom characteristics improved the screening efficiency of PLGC, but its effect was slightly lower than that of tongue image characteristics. In addition, there was a low consistency between tongue image characteristics and the appearance of gastric symptoms (Appendix 4). Therefore, we found that the introduction of symptom characteristics on top of tongue image characteristics had a small improvement in the effect of PLGC screening.

**Figure 5.**
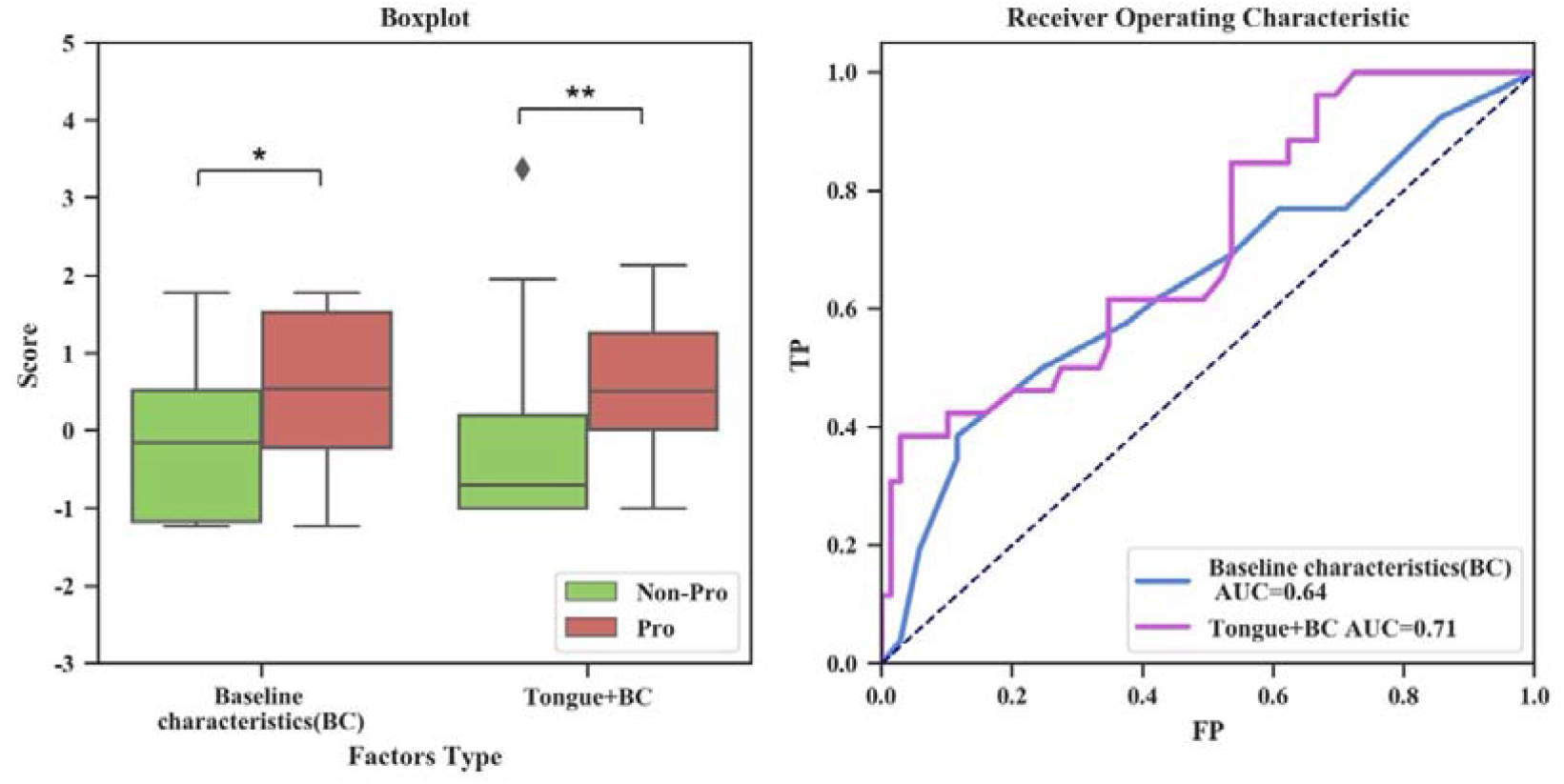
Boxplot of screening score and ROC curves comparisons with and without the inclusion of tongue image characteristics for risk prediction of PLGC. (**: p<0.01, **: p<0.05)

### Evaluation of the validity of tongue image characteristics for risk prediction of PLGC

We further resolved the ability of the model for risk predicting of PLGC by establishing the follow-up cohort, in which the initial pathological test was non-PLGC. Patients were divided into progressive (Pro) and non-progressive (non-Pro) groups with a second test as PLGC and non-PLGC (Table 1). Based on the previous logistic regression PLGC screening model based on the validation cohort, all patients in the Follow-up Cohort were scored for PLGC risk. The introduction of tongue image characteristics compared to baseline metrics showed a significant increase (p<0.01) in the differentiation of risk scores between the two groups, with an AUC increase of 10.9% (0.64 to 0.71) (Figure 5). In addition, we performed a univariate analysis of TDL for risk prediction of PLGC, and the results were in Appendix 5. Therefore, tongue image characteristics were potentially valuable for enhancing the predictive ability of PLGC risk, and further research was needed.

## Discussion

Screening patients with PLGC is important for the prevention and treatment of gastric cancer. In this study, we analyzed the tongue image characteristics associated with PLGC and based on this, constructed a PLGC screening model on a development cohort, then externally validated it in an independent validation cohort and evaluated the ability of risk prediction of PLGC in a follow-up cohort. Our study demonstrated the value of tongue image characteristics in PLGC screening and its potential for risk prediction.

We found that H. pylori infection was weakly correlated with PLGC and non-PLGC, although Hp infection is the most prominent risk factor for GC. Similar results have been found in other studies on the prediction of gastric cancer risk[47]. In this study, PLGC was analyzed with symptoms. We found only a small proportion of symptoms correlated with PLGC and their screening efficiency was not high, which is consistent with the findings of other studies [48].

Our proposed method has better performance than another study in screening of PLGC. Wang *et al*. developed a model with non-invasive indicators for PLGC screening including 290 patients with gastritis and the AUC was 0.728 (95% CI [0.651-0.793]) while our method was 0.76[49].

The study has limits. The data source was biased compared to the natural population. Due to the need for accurate information on the stage of gastritis, the data for establishing the system all came from patients with gastric disease, which had a certain deviation compared with the natural population. We have developed a smartphone-based App screening system to enhance the application convenience of the AITongue model in the natural population (Appendix 6). In further studies, more samples would be collected from natural populations to reduce bias and larger external validation should be done.

The screening model constructed in this study could improve the accuracy of PLGC screening. Tongue image characteristics were validated for their value in PLGC screening and risk prediction, which may drive tongue image characteristics as a new risk indicator in the future. By extracting tongue image characteristics through deep learning techniques, this study proposes a new approach for non-invasive PLGC screening and shows the possibility of its being used in large-scale applications.

## Data Availability

All data produced in the present study are available upon reasonable request to the authors

## Declarations

### Ethics approval and consent to participate

The experimental protocol was established, according to the ethical guidelines of the Helsinki Declaration and was approved by the Human Ethics Committee of Institution Review Board of Tsinghua University.

### Consent for publication

Not applicable.

### Competing Interests

The authors declare that they have no competing interests.

### Funding

Funding for this study was provided by the National Natural Science Foundation of China, China [81225025 and 62061160369]; and the Beijing National Research Center for Information Science and Technology, China [BNR2019TD01020 and BNR2019RC01012].

### Authors’ Contributions

Changzheng Ma contributed to the study design, data collection, data analyses, and writing. Peng Zhang, Xinxing Lai, Aidi Tan, and Xin Wang contributed to the study design and writing. Chaofan Ji, Qingrui Zhang, Shiyu Du, and Yan Li contributed to the data collection. Shao Li is the corresponding author.

## Acknowledgments

Thanks to the doctors and nurses of China-Japan Friendship Hospital and Yijishan Hospital of Wannan Medical College for their support of data collection.

## Notes

### Competing Interest Statement

The authors have declared no competing interest.

### Author Declarations

The experimental protocol was approved by the Human Ethics Committee of Institution Review Board of Tsinghua University.

