## Appendix 1-6 for "Construction of tongue image-based machine learning model for screening patients with gastric precancerous lesions"

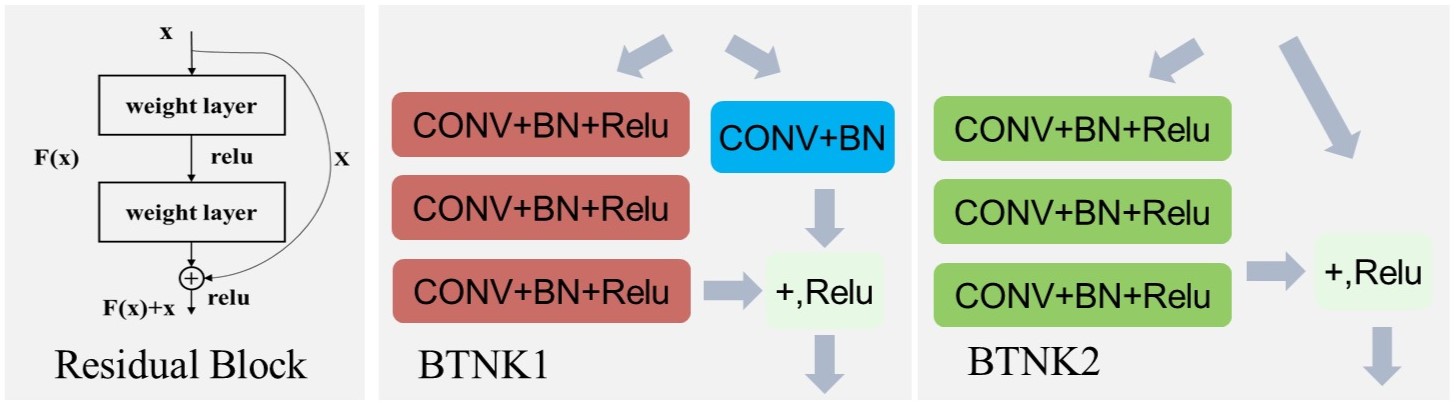

Appendix 1. The residual module of the ResNet50 model.

Appendix 2. Univariate and multivariate analysis of symptoms factors in PLGC screening.

| Variable | Total (n=1995) NO. (%) | non-PLGC (n=1824) No. (%) | PLGC (n=171) No. (%) | P value* | Adjusted OR (95% CI) † | P value |
| --- | --- | --- | --- | --- | --- | --- |
| **Symptoms Factors** | | | | | | |
| **Xerostomia** |  |  |  | 5.9e-6 |  |  |
| Yes | 642(0.32) | 560(0.31) | 82(0.48) |  | 2.07([1.41,3.04]) | 1.9e-4 |
| No | 1353(0.68) | 1264(0.69) | 89(0.52) |  |  |  |
| **Bitter Taste** |  |  |  | 2.7e-2 |  |  |
| Yes | 412(0.21) | 365(0.20) | 47(0.27) |  | 1.54([1.01,2.36]) | 4.7e-2 |
| No | 1583(0.79) | 1459(0.80) | 124(0.73) |  |  |  |
| **Belching** |  |  |  | 2.7e-2 |  |  |
| Yes | 518(0.26) | 461(0.25) | 57(0.33) |  | 1.72([1.15,2.56]) | 7.9e-3 |
| No | 1477(0.74) | 1363(0.75) | 114(0.67) |  |  |  |

*P values refer to the comparison between PLGC and non-PLGC groups in the univariate analysis.

†For variables not significant (p>0.05) in the logistic regression model, multivariable data are not shown.

Appendix 3. Univariate analysis of tongue diagnostic and symptoms factors in PLGC screening.

| Variable | Total (n=1995) No. (%) | non-PLGC (n=1824) No. (%) | PLGC (n=171) No. (%) | P value | OR (95% CI) |
| --- | --- | --- | --- | --- | --- |
| **Tongue Diagnostic Labels** | | | | | |
| **Teeth marks** |  |  |  | 0.49 |  |
| Yes | 347(0.17) | 321(0.18) | 26(0.15) |  | 0.84([0.54, 1.30]) |
| No | 1648(0.83) | 1503(0.82) | 145(0.85) |  |  |
| **Spotted** |  |  |  | 0.51 |  |
| Yes | 29(0.01)) | 28(0.02) | 1(0.01) |  | 0.38([0.06, 2.58]) |
| No | 1966(0.99) | 1796(0.98) | 170(0.99) |  |  |
| **Symptoms Factors** | | | | | |
| **Stomach dull pain** |  |  |  | 0.17 |  |
| Yes | 273(0.14) | 256(0.14) | 17(0.10) |  | 0.68([0.40, 1.13]) |
| No | 1722(0.86) | 1568(0.86) | 154(0.90) |  |  |
| **Constipated** |  |  |  | 0.20 |  |
| Yes | 104(0.05) | 91(0.05) | 13(0.08) |  | 1.57([0.86, 2.85]) |
| No | 1891(0.95) | 1733(0.95) | 158(0.92) |  |  |
| **Stomach bloating** |  |  |  | 0.33 |  |
| Yes | 1491(0.75) | 1369(0.75) | 122(0.71) |  | 0.83([0.58, 1.17]) |
| No | 504(0.25) | 455(0.25) | 49(0.29) |  |  |
| **Belching** |  |  |  | 0.38 |  |
| Yes | 1108(0.56) | 1019(0.56) | 89(0.52) |  | 0.86([0.63, 1.17]) |
| No | 887(0.44) | 805(0.44) | 82(0.48) |  |  |
| **Loose stools** |  |  |  | 0.60 |  |
| Yes | 53(0.03) | 50(0.03) | 3(0.02) |  | 0.63([0.20, 2.03]) |
| No | 1942(0.97) | 1774(0.97) | 168(0.98) |  |  |
| **Stomach stinging** |  |  |  | 0.60 |  |
| Yes | 66(0.03) | 62(0.03) | 4(0.02) |  | 0.68([0.25, 1.88]) |
| No | 1929(0.97) | 1762(0.97) | 167(0.98) |  |  |
| **Gastric cardialgia** |  |  |  | 1.0 |  |
| Yes | 38(0.02) | 35(0.02) | 3(0.02) |  | 0.91([0.28, 3.00]) |
| No | 1957(0.98) | 1789(0.98) | 168(0.98) |  |  |

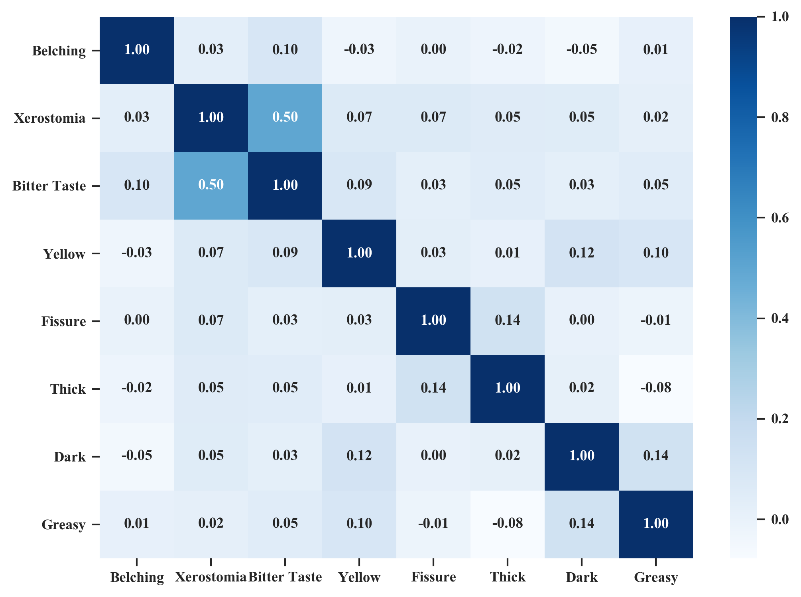

Appendix 4. Correlation analysis of TDL and symptoms.

Appendix 5. The univariate analysis of tongue diagnostic labels in risk prediction of PLGC.

| **Characteristics** | **Pro (26) Num (Rate)** | **non-Pro (69) Num (Rate)** | **OR** | **P value** |
| --- | --- | --- | --- | --- |
| **Coating (Yellow)** | 10(0.38) | 6(0.09) | 6.35 | 2.1e-3 |
| **Greasy** | 6(0.23) | 15(0.22) | 1.04 | 1.00 |
| **Fissure** | 5(0.19) | 8(0.12) | 1.76 | 0.56 |
| **Coating (Thick)** | 8(0.31) | 11(0.16) | 2.26 | 0.21 |
| **Dark** | 3(0.15) | 7(0.16) | 0.96 | 1.00 |

Appendix 6. The operation interface of the App.

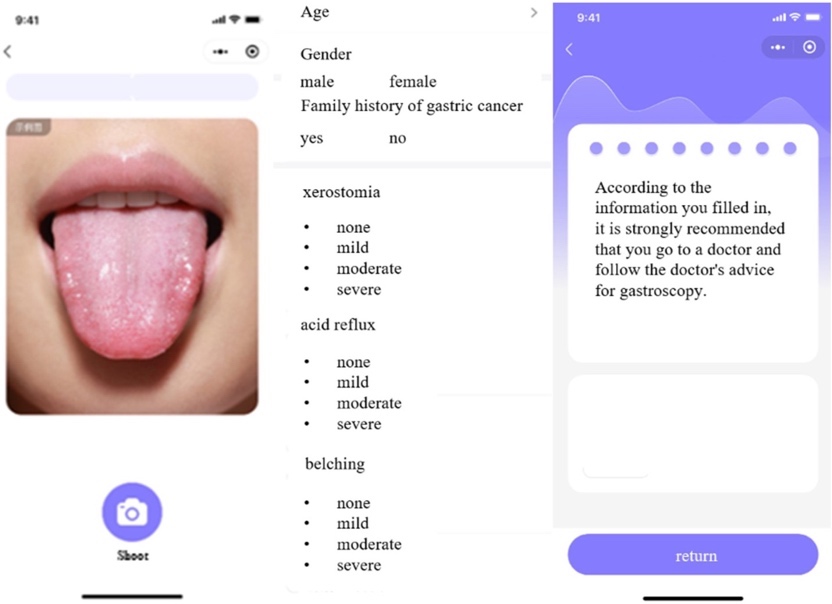
